# Implementation Fidelity of Tuberculosis Screening for Diabetes Mellitus Patients Among Healthcare Providers Offering Diabetes Mellitus Services at Public Health Facilities in Ubungo District, Dar es Salaam, Tanzania

**DOI:** 10.64898/2025.12.12.25342133

**Authors:** Edwin Christian Chavala, Felistar Mwakasungura, Linda Simon Paulo, Tumaini Nyamhanga

## Abstract

Globally, the risk of acquiring Tuberculosis (TB) among Diabetes mellitus (DM) patients is three (3) times higher than in the general population. Patients with DM not only have a high risk of getting TB disease but also have poor treatment outcomes Despite the National TB guideline recommending TB screening among DM patients, adherence remains low. This study aims to assess the implementation fidelity (IF) and factors affecting TB screening for DM patients among providers offering DM services in public health facilities in Ubungo district.

We conducted a descriptive cross-sectional study from April 4^th^ to May 25^th^, 2025, in 20 public facilities (3 hospitals, 5 health centers, 12 dispensaries) in Ubungo district using quantitative methods among 94 health providers offering DM services. Data were collected through a questionnaire and analyzed for fidelity levels (low or high) using descriptive statistics. Then, regression models using STATA version 16 identified factors affecting the fidelity of TB screening for DM patients among the healthcare providers offering DM services.

The overall fidelity score was 83.0% with (n=78) out of 94 providers achieving high fidelity, with only 17.0 % (n=16) of the total providers having lower fidelity levels. Teamwork (aPR 2.28, 95% CI 1.11–7.12; p-value =0.031), self-efficacy (aPR 2.29, 95% CI 1.04–5.02; p-value =0.024), and facility-level the provider was working, especially hospital level (aPR 3.60, 95% CI 1.52–8.50; p-value =0.004) were significantly associated with IF of TB screening for DM patients among healthcare providers.

Key factors influencing TB screening for DM patients among healthcare providers were effective teamwork, self-efficacy, and the level of the facility where the healthcare was working, such as hospitals. Therefore, strengthening teamwork and provider self-efficacy through training, is critical in universalizing high-fidelity practice and accelerating TB screening for DM patients among healthcare providers offering DM services in Ubungo district.

## Introduction

TB remains one of the deadliest infectious diseases worldwide, ranking as the second leading cause of death annually (1).According to the World Health Organization (WHO) 2022 report, the estimated global number of newly identified TB cases reached 7.5 million, surpassing the previous peak of 7.1 million reported in 2019 before the COVID-19 pandemic. Moreover, about one-quarter of the world’s population is infected with Mycobacterium tuberculosis, with a lifetime risk of progressing to active disease ranging between 5% and 10% (2).

People with weakened immune defenses or existing health conditions such as Diabetes Mellitus (DM) face a higher risk of developing TB (3). There is also no doubt that DM has recently been recognized as a major risk factor for TB, ranking among the top five contributors to TB burden. The sharp global rise in DM cases has highlighted the need to examine the intersection between DM and TB, positioning it as a key issue in public health. Research indicates that individuals with DM are about two to three times more likely to develop TB compared with the general population, and this increasing trend of DM may complicate the TB prevention and control efforts (4). More than half of the world’s TB cases occur in countries that have significant prevalence rates and total numbers of DM cases (5). Approximately 15% of patients with TB are found to have concurrent DM, a coexistence that has been identified as a potential catalyst for significant future public health challenges(6). In response to this, the World Health Organization, International Diabetes Federation (IDF), and International Union Against Tuberculosis and Lung Disease recommended routine TB screening for patients with DM (7,8).

In Tanzania, despite the national TB guideline recommending TB screening among DM patients, adherence remains low, as it was found to range from 1.3–14% which indicates that routine screening for TB among patients with DM is still often underdiagnosed (1,9,10). The dual burden of tuberculosis among individuals with diabetes in Tanzania has not yet received sufficient attention as an emerging public health concern at both local and national levels, with a 2012 nationwide survey of the working-age population reporting a DM prevalence of 9.1%, including those with elevated fasting blood glucose or already receiving diabetes treatment (11). Yet another study carried out in Tanzania revealed that TB prevalence among people with DM was seven times greater than the national average (7). Furthermore, findings in the Kagera region indicated that 14% of TB patients in Tanzania were also living with diabetes (1). Not only that, but also in a study that included Ubungo and other districts revealed that out of 619 facilities reported providing DM services, only 238 (38·4%) of them provided screening and treatment for TB. Therefore, we carried out this study to assess how consistently TB screening is carried out for DM patients and factors influencing its implementation among providers offering DM services in Ubungo District.

## Materials and Methods

### Study design and area

A descriptive cross-sectional study design employing a quantitative approach was used to collect and analyze data. The study was conducted in Ubungo District, located in Dar-es-Salaam Region. Ubungo is one of the five councils in the region, covering an area of 269.4 km². It shares borders with Kinondoni District and Kibaha (Pwani Region) to the north, Kisarawe (Pwani Region) to the west, and Ilala District to the south and east(12). Among the five districts, Ubungo reported a noticeable rise in cases of DM throughout the two years, with a substantial rise of 2808 cases from 2022 towards 2024, representing a 13.4% increase, leading to a total of 23,538 confirmed DM patients based on data from the DHIS2 accessed at 12:23 hours on 15^th^ January,2025 (13). Even with these high numbers, there is limited research on how well TB screening for DM patients is being carried out by the DM healthcare providers in Ubungo District.

### Study population and sampling

The study population included all clinicians providing DM services in Ubungo district, as they are the frontline staff responsible for daily screening of signs and symptoms, provision of patient education, counseling, and making referrals, according to Tanzania’s National TB Guideline for all eligible patients attending public health facilities. Their direct role in patient care across Ubungo District allowed the study to accurately assess how closely TB screening practices are followed as per the National TB guideline among DM providers. A finite population correction for proportion was used to calculate the sample size based on Cochran’s formula N=Z^2^ p(1-p)/e^2^, with a prevalence of adherence set at 13%, and margin of error set at 5%, and accounting for a 10% non-response rate. From each of the facilities, 2-4 participants were proportionately selected, making a total of 94 clinicians aged 18 and above from all public health facilities offering DM services in Ubungo District, with each selected facility contributing a sample proportionate to its total number of clinicians. Then, simple random sampling, using a random number generator, was applied to select providers until the required sample size was reached. The exclusion criterion was clinicians who held administrative roles without clinical responsibilities related to TB and DM services. To address potential missing data or non-responses, the scope of healthcare providers was widened; thus, additional healthcare providers were selected as potential replacements. This approach ensured that the final sample remained representative and that sufficient data were collected for accurate analysis.

### Variables and Measurements

Fidelity as the outcome of interest was measured using two constructs, which were details of content and frequency. Content were a Yes/No questions (1=yes activity done 0=No activity not done) while frequency referred to the extent to which clinicians at the facilities offering DM services performed TB screening activities on the guideline according to the prescribed frequency which was measured on a scale of 1 to 5 with a score ranging from 0 = never, 1 = rarely, 2 = for new clients 3 = For most clients 4=All the time for all clients), with a score 4 being what the intervention guidelines expected (14). We further re-grouped the original items selected from the two constructs and frequency into three domains, components which were TB signs and symptoms, TB awareness (education and counseling), and TB referrals based on our knowledge and experience. Each of the three domain components had different items that must be implemented to achieve the purpose of TB screening as per the National TB guideline. For each of the three components, individual item scores were summed to generate a domain-specific fidelity score. To standardize the scores and facilitate interpretation, percentage scores were calculated by dividing each of the three component scores by its maximum possible value and multiplying by 100. Then the overall fidelity score was computed by merging the three domain components. Based on the various scores for all three domain components, the total maximum possible score was 40, with 16-items per provider. TB symptoms and signs had a maximum score of 25; TB awareness (education & counseling) 10, and TB referrals had 5 a maximum score. To proceed with assessing the provider fidelity level, we conducted an internal consistency and reliability check to ensure that the various items are suitable to measure each of the three domain components. To evaluate the internal consistency of the grouped fidelity items within each domain component. The Cronbach’s alpha statistic measure for consistency was calculated and showed acceptable to excellent internal consistency for the scale components (15). TB signs and symptoms scored 0.70, TB education and counseling 0.97, while TB referral, assessed by a single item, did not require reliability testing. The factors affecting implementation fidelity of TB screening among providers offering DM services were system characteristics such as (training, teamwork, records documentation, staff allocation) while guideline characteristics included (Nature and source of the TB guideline, relative advantage and finally design quality and package of the TB guideline) which were measured based on a 5-point Likert scale, and were coded ranging from 1 for “strongly disagree” to 5 for “strongly agree” with 3 representing neutral position. Furthermore, provider characteristics such as the provider’s familiarity with TB screening for DM patients, as per the National TB guideline, and providers’ self-efficacy, which were measured and coded ranging from 1 for “very poor” to 5 for “very good”.

### Data collection tool and quality assurance

A self-administered structured questionnaire was adopted and modified based on a study from Ghana that measured implementation fidelity of TB screening among healthcare providers was used to collect data from participants (16). It gathered information on providers’ demographic characteristics, their reported adherence to clinical guidelines for TB screening among diabetes patients (covering content and frequency, as outlined in the TB/DM collaborative protocol in the National TB Guidelines), as well as factors influencing the fidelity of TB screening, based on the Conceptual Framework for Implementation Fidelity (CFIF) (17). Research assistants received three days of training, followed by a fourth day dedicated to pretesting. The pretest was conducted in a Mwananyamala hospital, Buguruni health center, and Bungoni dispensary that had similar characteristics to the study sites but were not part of the main study. This process was done to minimize measurement errors, reduce respondent burden, and identify any problem areas in the tools for correction(18). Data for this study were collected between April 4 ^th^ to 25^th^ May 2025 in 20 public health facilities that provide diabetes services, including 3 hospitals, 5 health centers, and 12 dispensaries across Ubungo District in Dar-es-Salaam Region.

### Data management and analysis

The computer program Epi Info version 3.5.1 software was used to capture the collected data based on a hardcopy questionnaire and further imported into Stata 16. To ensure data security, the file was saved in a password-protected location accessible only to the investigator. Before analysis, a thorough cleaning process was undertaken so as to remove missing numbers, outliers, and other irregularities, and this involved verifying the accuracy and completeness of the information collected from participants before analysis. To assess the level of implementation fidelity of TB screening for DM patients, normality tests using skewness were first performed using STATA version 16(Stata Corp, College Station, TX, USA) to determine data distribution. Summary statistics were then calculated for implementation fidelity using the two constructs of details of content and frequency, which were further merged into three domains which were TB signs and symptoms, TB awareness (education and counseling), and TB referrals to get scores among healthcare providers. Fidelity was categorized as either high (≥60%) or low (<60%), based on a threshold adopted from a prior study in Ethiopia (19). To examine factors affecting the implementation fidelity of TB screening for DM patients among clinicians, unadjusted and adjusted Poisson regression models were used (20). These models evaluated the relationship between fidelity and various system, provider, and guideline characteristics. This started with Model 1 assessed crude associations to determine individual association with fidelity to the National TB guideline, while Model 2 adjusted for potential confounders. Variables with an association that had a p-value < 0.2 were thereafter subjected to a multiple regression analysis to determine an independent association after controlling for confounders and covariates. Statistical significance was set at p<0.05 (21). The variance inflation factor (VIF) was computed to assess the presence of multicollinearity among the regression variables. For TB screening among DM patients, VIF values ranged from 1.08 to 1.47, with a mean VIF = 1.23, indicating no evidence of multicollinearity. All analyses were conducted using Stata version 16 (22).

### Ethical considerations

Permissions were granted by the public health facilities involved. Ethical clearance (Ref No. DA.282/298/01.C/2735) was obtained on 21^st^ March 2025 from the MUHAS ethical review committee. All participants were informed about the study aims, provided written consent, and were assured that participation was voluntary with the right to withdraw at any time. No incentives were offered. Confidentiality was maintained throughout by using identity numbers instead of names.

## Results

### Demographic information of study participants

A total of 94 healthcare providers participated in this study, of whom came from those offering DM services. Most of the providers offering DM services were aged 18–34 years 61 (64.9%) and slightly more than half were male 50 (53.2). The majority 54 (57.5%) had a diploma or below, and worked primarily at dispensaries 20 (21.3%), health centers 41 (43.6%). Most providers had one to five years of work experience 71 (75.5%). The mean age of the study participants was SD 37±6.6 years.

**Table 1:**
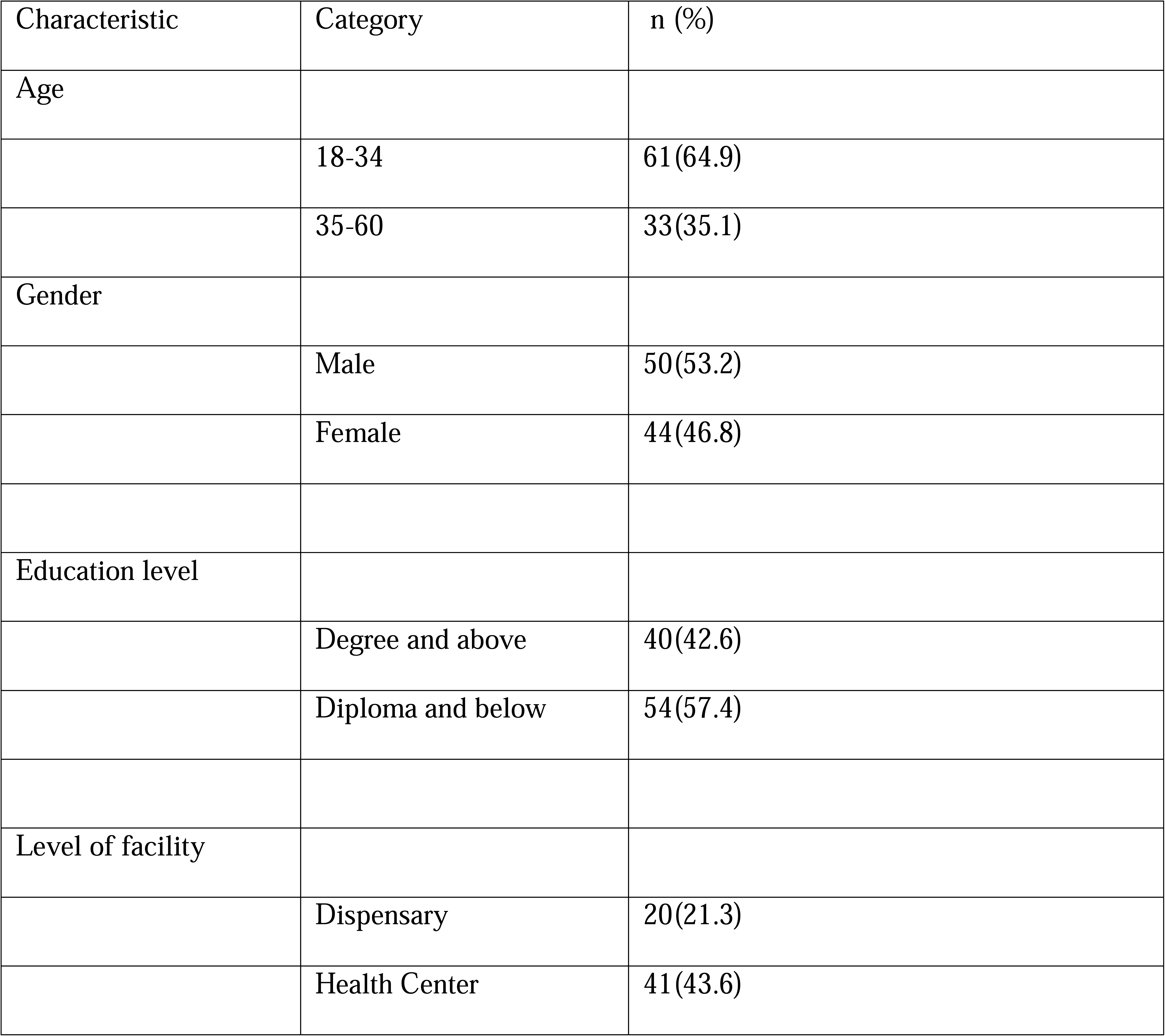
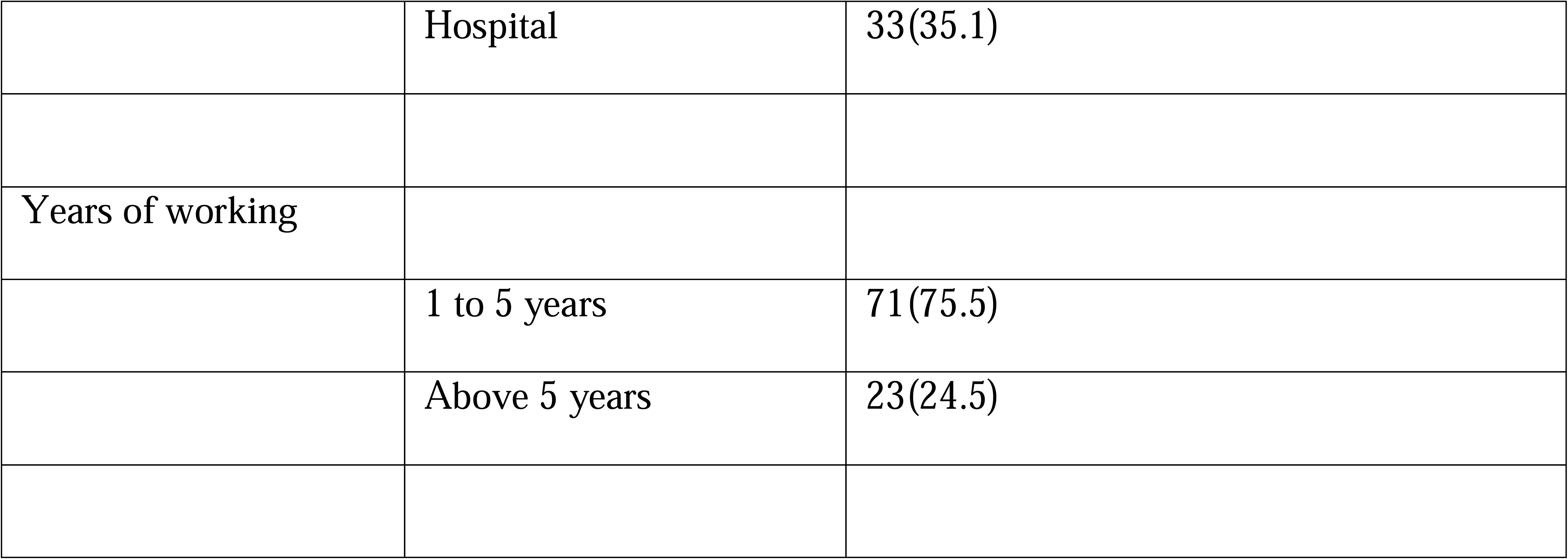
Social demographic characteristics of the healthcare providers offering DM services(n=94).

### Descriptive statistics on the implementation fidelity of TB screening for DM patients among providers offering DM services based on constructs of content and frequency (N=94)

The table below presents the descriptive statistics for the raw data on implementation fidelity for TB screening among providers offering DM services. For example, 72.3% (n=68) of DM providers give TB education to DM patients, while 93.6% (n=88) provide referrals for DM patients suspected of TB upon screening.

**Table 2:**
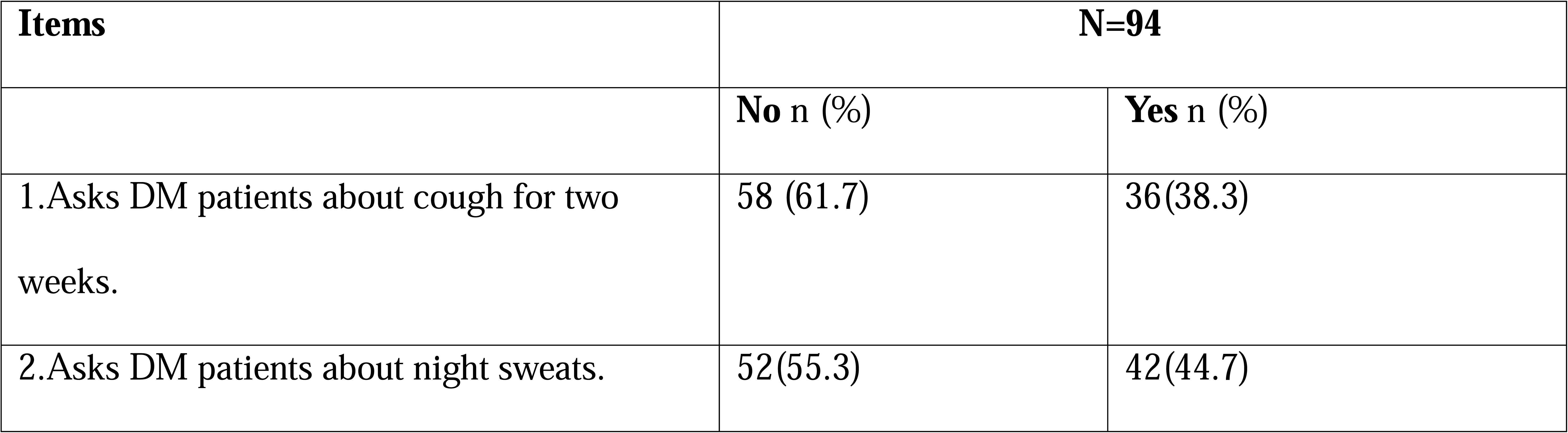
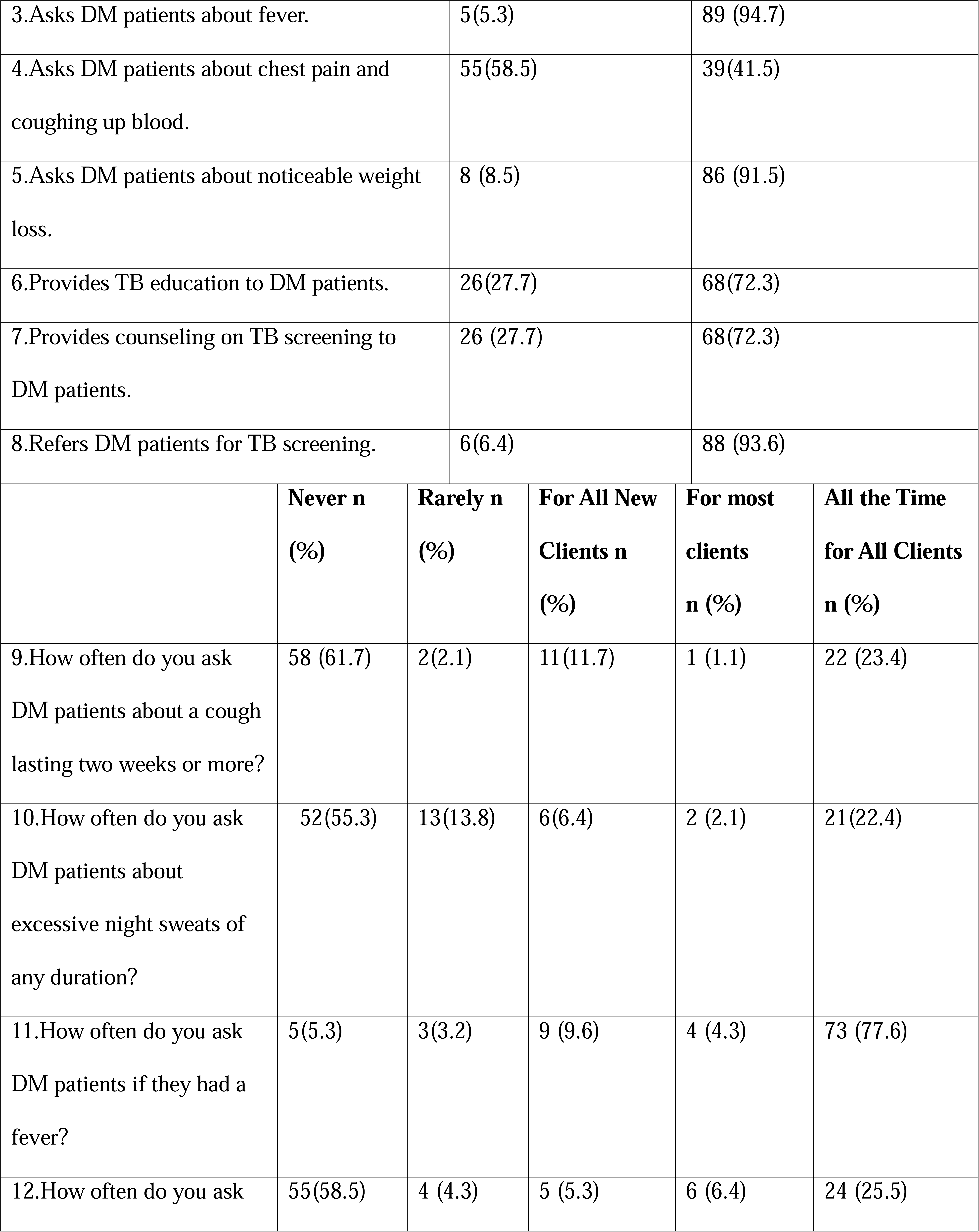
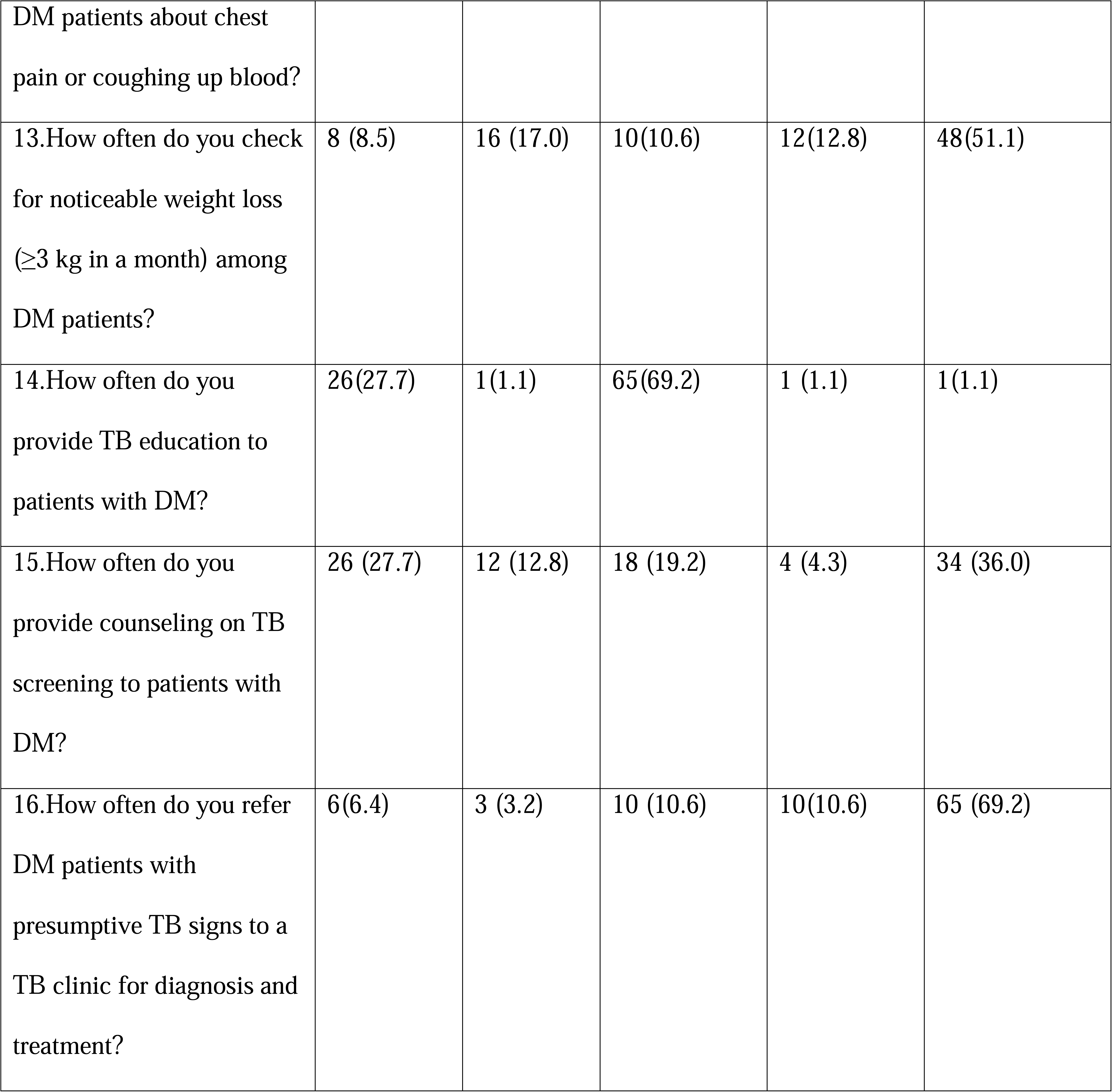
Descriptive statistics of the fidelity assessment items for TB screening among DM healthcare providers in public health facilities in Ubungo district.

The overall fidelity score of TB screening among providers delivering DM services was derived from three components: TB symptoms/signs, TB education and counseling, and TB referral across the two constructs of content and frequency.

### TB signs and symptoms

Out of 94 DM providers, 92.6% (n=87) had scores above the cut-off point score on asking about the TB signs /symptoms for DM patients.

### TB education and counseling

Moreover, on TB education and counselling among DM providers, out of 94 DM providers, 71.3% (n=67) had scores above the cut-off point on providing the TB education and counselling for DM patients.

### TB referrals

Lastly, provision of TB referrals for DM patients suspected of TB among the DM providers showed that out of 94 DM providers, 93.6% (n=88) had scores above the cut-off point on providing TB referrals for DM patients suspected of TB based on signs and symptoms.

Thus, the overall implementation fidelity score of TB screening for DM patients was higher (83.0%, n=78) out of the 94 providers offering DM services reported to screen TB for DM patients, reflecting a robust implementation fidelity, with only (17.0%, n=16) having a lower fidelity.

**Table 3:** Overall implementation fidelity of TB screening for DM patients among healthcare providers offering DM services. (n=94)

| Component | Low fidelity<br>n (%) | High fidelity<br>n (%) |
| --- | --- | --- |
| TB signs/symptoms | 7(7.4) | 87(92.6) |
| TB awareness<br>(education & | 27(28.7) | 67(71.3) |
| counseling) |  |  |
| awareness |  |  |
| TB referrals | 6(6.4) | 88(93.6) |
| Over all | 16(17.0) | 78(83.0) |

Moreover, the fidelity levels of TB screening according to the National TB guideline differed across various healthcare facilities. Hospitals and health centers showed the highest fidelity levels, with hospitals having 45.8%, and health centers 43.8% reflecting strong compliance with the guidelines. In contrast, dispensaries recorded the lowest adherence rate at 10.4%, indicating possible challenges in sustaining guideline compliance within these facilities (Figure. 1).

**Figure 1:**
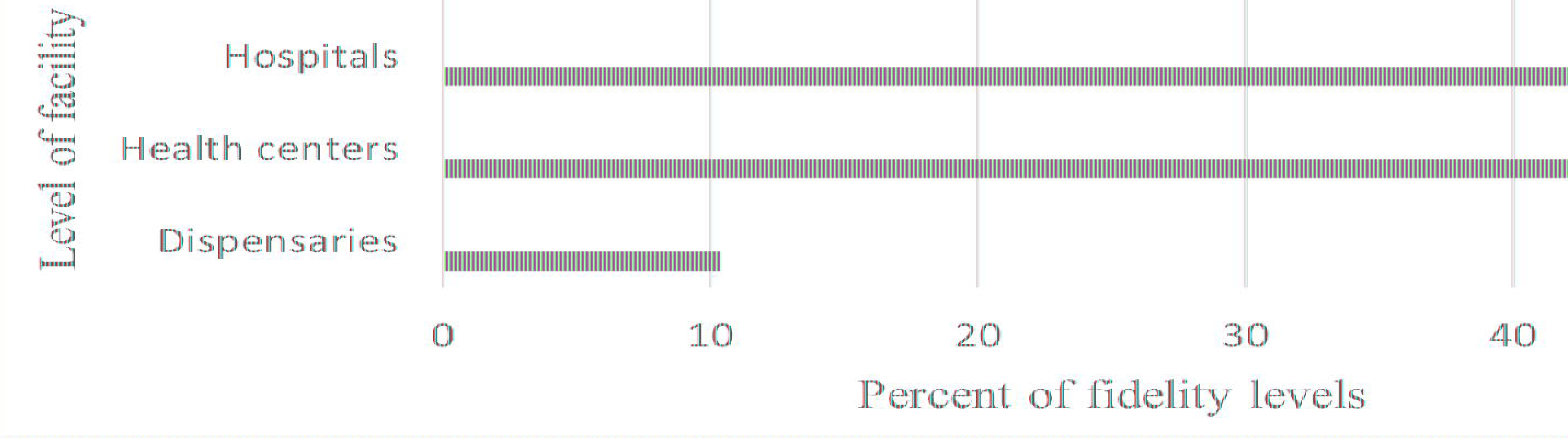
Fidelity levels among the level of facility.

### Factors associated with provider implementation fidelity of TB screening for DM patients among healthcare providers offering DM services

We used the modified poison regression on system-related factors, provider characteristics, Guideline characteristics, and social demographics characteristics to identify factors affecting the implementation fidelity of TB screening among providers offering DM services. After adjusting for other factors among the system characteristics, teamwork among staff (aPR=2.28195% CI: 1.11-7.12; p-value =0.031) was significantly associated with higher fidelity; moreover, also among the providers’ characteristics, self-efficacy (aPR=2.29; 95% CI: 1.04-5.02; p-value 0.024) was significantly associated with higher fidelity. This suggests that DM health providers with strong teamwork were twice as likely to conduct TB screening, while those with self-efficacy were twice as likely to adhere to TB screening protocols for DM patients. While training showed associations in the crude model, they did not retain statistical significance in the adjusted analysis. Regarding the social demographics of study participants, the level of the facility was strongly associated with fidelity.DM providers at hospitals were three times more likely (aPR=3.60, 95% CI 1.52-8.5; p-value 0.004) to adhere to TB screening for DM patients with higher fidelity compared to those at health centers and dispensaries.

**Table 4:** Factors.

| Variable | Crude PR (95%CI) | p-value | Adjusted aPR(95%CI) | p-value |
| --- | --- | --- | --- | --- |
| Self-efficacy | 2.40(1.22-4.67) | <0.01 | 2.29(1.04-5.02) | 0.024 |
| Familiarity to TB guideline | 1.66(1.13- 2.43) | <0.01 | 1.19(0.73- 1.93) | 0.480 |
| Nature and source of TB guidelines | 1.16(0.54- 2.50) | 0.695 | 0.79(0.40- 1.55) | 0.493 |
| Relative advantage | 1.01(0.52- 1.95) | 0.972 | 1.55(0.64- 3.75) | 0.332 |
| Design quality | 1.40(0.72- 2.75) | 0.322 | 1.43(0.67- 3.07) | 0.355 |
| Teamwork among staff | 2.08(1.23-4.97) | <0.01 | 2.28(1.11-7.12) | 0.031 |
| Training | 0.62(0.40-0.94) | 0.024 | 0.63(0.38-1.09) | 0.104 |
| staffing | 0.91(0.49- 1.70) | 0.769 | 1.03(0.43- 2.48) | 0.526 |
| Social demographics characteristics |  |  |  |  |
| Age |  |  |  |  |
| 18-34 | Ref |  |  |  |
| 35-60 | 1.21(0.81-1.79) | 0.343 | 1.089(0.72-1.63) | 0.678 |
| Gender |  |  |  |  |
| Female | Ref |  |  |  |
| Male | 0.88(0.59-1.30) | 0.528 | 0.86(0.58-1.23) | 0.485 |
| Education level |  |  |  |  |
| Degree and above | Ref |  |  |  |
| Diploma and below | 0.97(0.64-1.44) | 0.532 | 2.21(0.93-5.27) | 0.061 |
| Level of facility |  |  |  |  |
| Dispensary | Ref |  |  |  |
| Health Center | 0.20(0.12-0.14) | <0.01 | 2.21(0.93-5.27) | 0.072 |
| Hospital | 1.51(1.13-2.02) | 0.034 | 3.60(1.52-8.5) | 0.004 |
| Years of working |  |  |  |  |
| 1 to 5 years | Ref |  |  |  |
| Above 5 years | 0.97 (0.62-1.53) | 0.902 | 0.42 (0.12- 1.44) | 0.959 |

## Discussion

This study aimed to assess how providers implement TB screening among those delivering diabetes services in Ubungo District, as well as the factors influencing the level of implementation fidelity. While research on implementation fidelity in health is increasing, this study is among the few in our setting to examine fidelity as an implementation outcome for TB screening by DM providers in line with the National TB Guidelines in public health facilities. In contrast, most studies in Sub-Saharan Africa and our setting have focused mainly on intervention outcomes such as cure rates, deaths, or treatment completion (23,24).

From the structured questionnaires, the first key finding showed that the overall implementation fidelity score of TB screening among DM patients was high, with 83.0% of providers reporting that they screened DM patients for TB, indicating strong fidelity to implementation. However, 17.0% of providers demonstrated low fidelity in TB screening, suggesting possible challenges with adherence due to sub-optimal coverage due to lack of integrated TB-DM trainings among providers. Evidence from published literature indicates that such provider non-adherence or low fidelity to healthcare interventions among healthcare providers can reduce the likelihood of achieving desired outcomes and may render otherwise effective interventions less impactful in real-world practice. Furthermore, the study revealed that providers with a diploma or below (58.3%) demonstrated higher fidelity compared to degree-holders (41.7%), but also those in hospitals had a higher fidelity (45.8%) compared to those in health centers (43.8%) and dispensaries (10.4%). These findings are in agreement with other studies in other low- and middle-income countries (9) which reported high fidelity for TB screening in DM clinics in urban hospitals, and this is explained by the fact that urban hospitals benefited from better resources, trained staff, and oversight from National TB programs. Our study findings also align with a study in the Republic of the Marshall Islands that reported higher fidelity of TB screening among DM patients due to effective training and engagement policies that encouraged providers to take the course of action as recommended by the guidelines (25). Moreover, our study contrasts with a study in Malawi (26), which reported less consistent fidelity compared to our settings, and this justification can likely be explained due to sufficient awareness and familiarity with bidirectional TB screening protocols among healthcare providers in our settings compared to Malawi. Similarly, our study findings contrast with another study in Malawi (27) which reported a fidelity which was substantially lower than our study’s fidelity level and this, the disparity in fidelity levels can be explained by the fact that our settings may have better resource availability, more consistent training, or stronger integration into routine care, allowing for closer adherence to protocols and higher coverage compared to Malawi which may have resource-constrained healthcare environment, which likely posed greater operational challenges. With regards to higher fidelity among diploma and below level providers in our study was surprising, but could be explained by their greater involvement in direct patient care compared to degree-holders, who may focus on administrative roles. The findings highlight that high implementation fidelity for TB screening in DM patients is achievable, particularly in settings with better resources and training, as seen in urban hospitals. Therefore, to enhance TB screening fidelity in DM patients, future programs and research should prioritize consistent provider training so as to improve the low fidelity by enhancing their understanding to ensure that providers in low-level facilities also consistently adhere to TB screening protocols for DM patients.

The next key finding of our study are the contextual factors such as (system-related, provider-related, and Guideline characteristics) affecting the level of implementation fidelity to TB screening. The findings from our study showed that among the system characteristics, teamwork was significantly associated with higher implementation fidelity of TB screening for DM patients among health care providers offering DM services. These findings align with a study in Malawi and the USA (28) which found fidelity of TB screening among DM patients was associated with strong teamwork, and the possible justification could be due to task-shifting among health workers in DM clinics, that enabled them to share tasks among themselves, using everyone’s skills and support to follow TB screening steps for DM patients more accurately, and hence improving the quality of screening. Moreover, our study findings showed that among provider characteristics, self-efficacy and the level of facility a provider was working, such as at the hospital level, were significantly associated with higher fidelity of TB screening for DM patients. These findings in our study are supported by findings in Eswatini and Iran (63,79), which had similar results, and this is explained by the fact that providers who had individual belief in their own capabilities to execute courses of action to TB screening among DM patients, as those with greater exposure to training, reported better adherence. The findings highlight the need for further research, and program interventions should focus on fostering teamwork and self-efficacy through training to improve TB screening fidelity for DM patients among providers offering DM services in lower facilities, such as health centers and dispensaries, to alleviate this disparity in adherence level.

### Strengths and limitations of the study

The study explores TB screening among DM patients in Ubungo public health facilities. It provides insights to improve adherence to national TB guidelines. Data were collected from multiple facility levels to reflect service variations. Limitations included possible response bias from self-administered questionnaires. Additionally; the findings may have limited generalizability to private facilities or regions with different resource levels.

## Conclusion

Our findings showed that achieving high implementation fidelity was feasible, implying that effective teamwork, provider self-efficacy, and the type of facility the provider was working such as those in hospital-level were associated with higher fidelity in TB screening. However, the low fidelity observed among some providers highlights key challenges, including the absence of integrated TB-DM training and inconsistent team coordination, which undermine TB screening efforts. To improve fidelity in Ubungo district with regard to the above-mentioned challenges, strategies should include integrating TB and DM training with follow-up supervisory visits to reinforce adherence as per the national TB Guideline, but also displaying visual aids such as posters, checklists, and flowcharts of TB screening questions in public health facilities offering DM services. In addition, future research should adopt mixed-method approaches across different health facility types, including both public and private, with emphasis on qualitative and observational methods, rather than relying solely on structured questionnaires, to capture a more comprehensive assessment of the implementation fidelity of TB screening for DM patients.

## Data Availability

All data will be available upon request and following the stipulated Tanzanian policies on data sharing, data may be made available to researchers requesting access.

## Acknowledgement

The authors wish to sincerely thank everyone who contributed to making this study possible, including the Ubungo district authorities. We are also grateful to all study participants for their cooperation, which greatly contributed to the successful completion of this research. This study was undertaken as part of a Master’s degree in Project Management, Monitoring, and Evaluation in Health at Muhimbili University of Health and Allied Sciences.

